# Targeting a highly repetitive genomic sequence for sensitive and specific molecular detection of the filarial parasite *Mansonella perstans* from human blood and mosquitoes

**DOI:** 10.1101/2022.06.29.22277064

**Authors:** Nils Pilotte, Tamara Thomas, Michael Zulch, Allison R. Sirois, Corrado Minetti, Lisa Reimer, Steven A. Williams, Lori Saunders

## Abstract

**Background:** *Mansonella perstans* is among the most neglected of the neglected tropical diseases, and is believed to cause more human infections than any other filarial pathogen in Africa. Based largely upon assumptions of limited infection-associated morbidity, this pathogen remains understudied, and many basic questions pertaining to its pathogenicity, distribution, prevalence, and vector-host relationships remain unanswered. However, in recent years, mounting evidence of the potential for increased *Mansonella* infection-associated disease has sparked a renewal in research interest. This, in turn, has produced a need for improved diagnostics, capable of providing more accurate pictures of infection prevalence, pathogen distribution, and vector-host interactions.

**Methodology/Principal Findings:** Utilizing a previously described pipeline for the discovery of optimal molecular diagnostic targets, we identified a repetitive DNA sequence, and developed a corresponding assay, which allows for the sensitive and species-specific identification of *M. perstans* in human blood samples. Testing also demonstrated the ability to utilize this assay for the detection of *M. perstans* in field-collected mosquito samples. When testing both sample types, our repeat-targeting index assay outperformed a ribosomal sequence-targeting reference assay, facilitating the identification of additional *M. perstans-*positive samples falsely characterized as “negative” using the less sensitive detection method.

**Conclusions/Significance:** Through the development of an assay based upon the systematic identification of an optimal DNA target sequence, our novel diagnostic assay will provide programmatic efforts with a sensitive and specific testing platform that is capable of accurately mapping *M. perstans* infection and determining prevalence. Furthermore, with the added ability to identify the presence of *M. perstans* in mosquito samples, this assay will help to define our knowledge of the relationships that exist between this pathogen and the various geographically relevant mosquito species, which have been surmised to represent potential secondary vectors under certain conditions. Detection of *M. perstans* in mosquitoes will also demonstrate proof-of-concept for the mosquito-based monitoring of filarial pathogens not vectored primarily by mosquitoes, an approach expanding opportunities for integrated surveillance.

**Author Summary:** Infection with *Mansonella perstans* remains exceedingly common in many of the world’s tropical and sub-tropical regions. However, *M. perstans* is largely understudied due to the long-held belief that this pathogen is of little clinical significance. However, in recent years, many within the research community have begun to advocate for the increased study of this pathogen, pointing to evidence of mansonellosis-associated disease morbidity, the potential for *Mansonella* spp. to confound diagnostic testing for other pathogens, and the possibility for co-infections with *M. perstans* to impact disease progression and treatment outcomes for other infections. As a result of this growing appreciation of the importance of *M. perstans*, there exists a need for improved diagnostic options, capable of providing researchers with the tools required to accurately and effectively map infection, explore the pathogen-vector relationship, and determine pathogen prevalence. In response to these needs, we have developed a novel real-time PCR assay targeting a highly repetitive DNA sequence within the *M. perstans* genome. This index assay outperformed a ribosomal-sequence targeting reference assay when comparatively testing both human blood and field-collected mosquito samples. As such, this assay will provide researchers with an improved tool for the identification of *M. perstans* as part of various operational research efforts.

## Introduction

Responsible for documented human infection in over 30 sub-Saharan African countries as well as parts of Central and South America, *Mansonella perstans* is a widespread pathogen of the world’s tropical and sub-tropical regions [1]. Yet, despite a high prevalence of infection, and its presumed status as the most frequent human filarial parasite in Africa [2], *Mansonella* is under studied when compared with other human-infecting filarial pathogens. Marginal research interest and a lack of funding are direct results of the long-held belief that *Mansonella* is of limited clinical significance [1,3]. This has led to the dubbing of mansonellosis as among the most neglected of the neglected tropical diseases (NTD)s [1]. However, with few studies in the scientific literature aiming to assess the health impacts of *M. perstans* infection, and evidence from some endemic regions suggesting that up to 10% of *Mansonella-*infected individuals may suffer severe morbidities [4], a growing faction of the research community is championing the need for a more thorough investigation of the public health implications of widespread mansonellosis [1,5]. Concerns also exist that *M. perstans* infection could be indirectly impacting human health by confounding the results of serological-based assays or rapid diagnostic tests (RDTs) intended to diagnose closely-related filarial pathogens [6]. While the extent of such impacts remains uncertain, and a limited body of evidence has suggested that such cross-reactivity may be unlikely [7], this possibility has raised concerns within the lymphatic filariasis community, where the cross-reactivity of RDTs with filarial pathogens such as *Loa loa* is believed capable of significantly skewing survey results, impacting interpretations of intervention outcomes, and complicating disease mapping efforts [8,9].

Critical to any assessment of disease morbidity is the ability to sensitively and specifically identify the presence of the disease’s causative agent. Appropriate and sufficient diagnostic strategies are also essential for overcoming the challenges associated with infection mapping, assessment of prevalence, and impact modeling. Molecular methods facilitating vector incrimination are equally important, as a fundamental understanding of the vectoring capacity of potential arthropod hosts is critical to an understanding of any vector-borne pathogen’s transmission dynamics and capacity for spread. In the case of *M. perstans*, only a partial and insufficient understanding of the vector-pathogen relationship exists. While *Culicoides* spp. are the only proven vectors for *M. perstans* [3], the capacity for black flies of the genus *Simulium* [10–12] and at least one species of midge belonging to the genus *Leptoconops* to vector *Mansonella ozzardi*, a closely-related species, has been documented [13]. These findings, in conjunction with field studies reporting gross inconsistencies between vector and host prevalence levels for *M. perstans* [14,15], have led some within the community to theorize that mosquitoes might represent competent *M. perstans* vectors in certain settings or under certain conditions [2]. Historical studies demonstrating the partial development of *Mansonella* in both *Anopheles* spp. and *Aedes aegypti* mosquitoes lend further support to this possibility [10]. Since the spread and distribution of vector-borne diseases cannot be fully understood without a comprehensive understanding of the underlying vector-pathogen relationships, improved methodologies for incrimination are critical to all assessments of disease-related risks. Considering such dependencies, and recognizing the uncertainties surrounding the health impacts and programmatic significance of *M. perstans*, the development of diagnostic strategies capable of providing answers to the many open questions surrounding this pathogen should be prioritized by the research community.

Attempting to address this diagnostic need, we now describe the development of a novel, real-time PCR (qPCR) assay targeting a highly repetitive sequence within the *M. perstans* genome. Utilizing our previously described bioinformatics-based pipeline for assay development [16–18], we have created an assay that demonstrates improved analytical and clinical sensitivity versus a ribosomal sequence-targeting reference assay. Of note, we have demonstrated improved *M. perstans* detection when testing both human blood samples and mosquito pools for the presence of pathogen. While blood testing remains the most obvious use for a *M. perstans* assay, the utility of this assay for mosquito surveillance should be of considerable interest to the research community. In addition to its possible implications for purposes of vector incrimination, mosquito-based detection will serve to facilitate expanded *M. perstans* mapping efforts by allowing for integrated surveillance with other mosquito-borne pathogens. With a resurgence of interest in the possible uses of molecular xenomonitoring (MX) within the LF community [19–21] as well as other NTD communities [22–24], prospects for joint surveillance are expanding. Recognizing this opportunity, many donors and advocates have begun to champion integration strategies that serve to maximize investment through the creative expansion of testing [25]. The surveillance of hematophagic insects for both competently- and incompetently-vectored pathogens presents one attractive possibility.

## Methods

### Ethics Statement

Human blood sample collections occurred under approvals gained, and fully described, as part of a previously conducted study [26]. Briefly, ethical approval was obtained by the Liverpool School of Tropical Medicine Ethics Committee (<u>Research Protocol 17-035 A vector excreta surveillance system VESS to support the rapid detection of vector-borne diseases</u>) and the Council for Scientific and Industrial Research, Accra, Ghana. For all participants, written consent was provided by the participant, or by the participant’s guardian.

### Isolation of *M. perstans* Microfilariae

In order to isolate *M. perstans* microfilariae (mf) for downstream sequencing, approximately 250 µL of a banked venous blood sample, previously collected from a single patient as part of an unrelated study [27] underwent syringe-based filtration. To accomplish this, blood was first diluted 1:10 in 1 X PBS. Diluted blood was then passed through a 5 µM polycarbonate membrane filter (Sterlitech, Kent, WA) using a sterile syringe. Positive pressure allowed blood components and cells to pass through the filter, while causing the larger microfilariae to remain on the filter’s surface. Following filtration, 10 sample volumes (5 mL) of 1 X PBS were added to the syringe and passed through the filter, helping to rinse away residual blood components. The filter was then placed into a 15 mL conical tube, and 1 mL of 1 X PBS was repeatedly washed across the filter’s surface to free/dislodge the isolated microfilariae. The filter was then removed and the tube was briefly spun at 1,000 x g to pellet the collected microfilariae.

### DNA Extraction from *M. perstans* Microfilariae

Pelleted *M. perstans* microfilariae underwent DNA extraction using the DNeasy Blood and Tissue Kit (Qiagen, Germantown, MD). Extractions occurred in accordance with the manufacturer’s suggested protocol for the spin-column-based purification of total DNA from the blood or cells of animals with minor modifications. Briefly, microfilariae were resuspended in 200 µL of 1X PBS, followed by the addition of 20 µL of Proteinase K and 200 µL of Buffer AL. The sample was vortexed, incubated at 56 °C for 10 min, and 200 µL of 100 % ethanol was added. The sample was vortexed, transferred to a DNeasy Mini spin column, and spun at 6,000 x g for 1 min. Flow-through was discarded and the sample was sequentially washed with Buffers AW1 and AW2 in accordance with the manufacturer’s recommendations. DNA was then eluted from the spin column in 100 µL of Buffer AE via centrifugation at the recommended time and speed. To maximize DNA recovery, the elution product was subsequently reloaded into the spin column and passed through a second time via repeat centrifugation.

### Library Construction and Next-Generation Sequencing

In preparation for next-generation sequencing (NGS) a library was created using the Nextera DNA Flex Library Preparation Kit (Illumina, San Diego, CA). Using 60 ng of gDNA (at a concentration of 2ng/µL), a dual-indexed library was generated in accordance with the manufacturer’s suggested protocol. Following preparation, quantification, and purification, 250 µL of this library, diluted to a 10 pM concentration, was mixed with 7.5 µL of a 10 pM PhiX library to increase diversity. Paired-end sequencing then occurred on the MiSeq instrument (Illumina) using a 150-cycle v3 Reagent Kit (Illumina).

### Repeat Analysis

Raw sequencing reads were prepared for downstream analysis using Galaxy-based tools for pre-processing. To ensure that only high quality input reads were included, reads whose component bases failed to meet or exceed a quality score threshold of 10 at 95% or more of the read’s total base positions were excluded. A random subset of 500,000 interlaced reads, having passed filer, was then selected for analysis. To identify highly repetitive elements within the *M. perstans* genome, reads were analyzed using both the RepeatExplorer2 [28] and TAREAN [29] applications as previously described [16–18,30,31]. These analyses resulted in the clustering of like sequences, and identified repetitive DNA elements predicted to be highly represented within the *M. perstans* genome. Consensus sequences, generated for each repeat cluster and predicted to represent the most abundant versions of each respective parent repeat, were then selected for further analysis.

To validate the origins of candidate consensus sequences, each sequence was analyzed using the “Nucleotide BLAST tool” available from the National Center for Biotechnology Information (NCBI) website (https://blast.ncbi.nlm.nih.gov/Blast.cgi). Because the filtration process used to separate juvenile worms from host blood was imperfect, the resulting DNA extract contained both *M. perstans-* derived and human-derived sequences. Clusters were therefore searched for BLAST similarity to human sequences and those mapping to elements of the human genome were removed from consideration. From the remaining sequence elements, a candidate predicted to be of greatest prevalence within the collection of raw reads was selected as a possible assay target. This *M. perstans* target was named *Mansonella perstans* Repeat 1 (*Mp*R1) and will be referred to as such throughout this manuscript.

### Primer and Probe Design

Utilizing the default parameters of PrimerQuest Tool (Integrated DNA Technologies, Coralville, IA), a qPCR assay was designed for the amplification of the *Mp*R1 repeat. Following design, forward and reverse primers were synthesized, as was a probe labeled with a 6FAM fluorophore and double-quenched using ZEN/3IABkFQ chemistries.

### Assay Optimization and Validation

#### Primer optimization

In order to determine primer concentrations that would allow for optimal amplification, a series of reactions were prepared in which candidate forward and reverse primers were diluted to concentrations of 300 nM, 500 nM, and 800 nM. Reactions containing each concentration of forward primer were prepared in combination with each concentration of reverse primer, such that a 3 × 3 reaction matrix was created. All reactions were run using 10 µL of TaqPath ProAmp Master Mix (ThermoFisher Scientific, Waltham, MA), and probe at a 125 nM concentration. Reactions were run in 20 µL volumes containing 100 pg of *M. perstans* gDNA template. Cycling conditions consisted of an initial 22 min hold at 50 °C, followed by a 10 min hold at 95 °C. Following these incubations, 40 cycles of 95 °C for 15 sec, followed by 60 °C for 1 min occurred. Twelve replicate reactions were prepared for each combination of primer concentrations and mean Cq values were calculated from the results produced using each primer combination.

#### Determination of Optimal Reaction Temperatures

To identify the annealing/extension temperature capable of producing the lowest Cq values, a series of reactions were run employing a temperature gradient from 55 °C to 60 °C. Four replicate reactions were run utilizing each annealing/extension temperature in combination with optimal primer concentrations, and employing all other reaction parameters, and reagent and template concentrations described above. Following reaction completion, mean Cq values were calculated for the results generated at each temperature.

#### Examination of Analytical Specificity

To assess the specificity of our candidate primer/probe pairing, NCBI’s Primer-BLAST tool (https://www.ncbi.nlm.nih.gov/tools/primer-blast/index.cgi) was used to screen for predicted amplification against a broad range of assay targets. Following *in silico* analyses, using optimal primer concentrations and cycling temperatures as determined above, a specificity panel containing gDNA extracts isolated from the filarial parasites *Brugia malayi, Brugia pahangi, Loa loa, Acanthocheilonema viteae, Onchocerca volvulus, Wuchereria bancrofti, Dirofilaria immitis*, and *Mansonella ozzardi*, as well as human gDNA was tested. All extracts were used as template in triplicate 20 µL reactions with 100 pg of gDNA added to each reaction well.

#### Examination of Analytical Sensitivity

To determine the analytical limits of detection for our assay, an aliquot of *M. perstans* gDNA was 10-fold serial diluted, generating stocks at concentrations ranging from 100 pg/µL to 1 ag/µL. Utilizing optimal reaction conditions, all concentrations of template were tested in triplicate 20 µL reactions, with 1 µL of template added to each reaction well.

### Assessment of Reaction Efficiency

#### Creation of a positive control plasmid

Utilizing our qPCR primers, the assay’s target sequence was amplified from *M. perstans* pure gDNA by conventional PCR as described previously [17]. PCR products were then cloned into the pCR-Blunt II-TOPO vector (ThermoFisher Scientific) in accordance with the manufacturer’s suggested protocol. Following cloning reactions, transformations of NEB Express Competent *E. coli* (New England Biolabs, Ipswich, MA) were performed and transformation products were plated on selective media in accordance with published methods [17]. *E. coli* was then allowed to grow overnight, after which individual colonies were sampled by scraping with a pipette tip. Colonies containing plasmid with only a single copy of the *Mp*R1 repeat sequence were then identified using a combination of colony PCR and sequencing as previously described [17]. A stock of this plasmid, containing a single copy of the *Mp*R1 repeat, was used as a positive control for all future experiments.

#### Determination of reaction efficiency

In order to assess reaction efficiency, a preparation of positive control plasmid was titrated, undergoing dilutions to create stocks at concentrations ranging from 500 pg/µL to 1 pg/µL. Because the *Mp*R1-contianing plasmid is 3,644 bp in size, and the average mass of a nucleotide base pair is estimated to be 650 Da, 100 ag of plasmid was estimated to contain approximately 25 plasmid copies. From this number, estimated copy numbers were then calculated for each concentration of plasmid utilized in the dilution series. Utilizing optimized reaction conditions, nine to 11 replicate reactions were run with each plasmid concentration as template. A calculation of reaction efficiency was then performed.

### Assessment of Clinical Sensitivity

#### Comparative analysis of field-collected blood and mosquito samples

In order to assess the clinical sensitivity of our newly described repetitive sequence-targeting assay, DNA from 158 dried human blood samples, and 316 individual mosquito samples, previously collected and extracted as part of an unrelated study [26], were tested for the presence of *M. perstans* using both our index assay, and a previously described, ribosomal sequence-targeting reference assay [5,26]. All samples were tested using each assay’s respective optimal conditions, and testing was performed in duplicate. For blood sample testing, samples which produced inconsistent results during initial testing with either assay, were retested with the appropriate assay, again in duplicate, and were scored as positive if at least one of two retest replicates produced a positive result. In the event that both retest replicates failed to produce a detectable product, the sample was recorded as a negative. Due to limited samples volumes, mosquito samples which produced inconsistent results for either assay could not be re-tested, and were therefore eliminated from the study.

## Results

### Repeat Analysis and Assay Design

Following the removal of clustered sequences mapping to human DNA, the remaining repetitive sequence, of greatest representation within the prepared library, was selected as a potential assay target. The consensus sequence for this, the *Mp*R1 repeat, with a monomer unit predicted to be 281 base pairs in length, was utilized as a template for the candidate assay’s primer and probe design (Table 1).

**Table 1.**
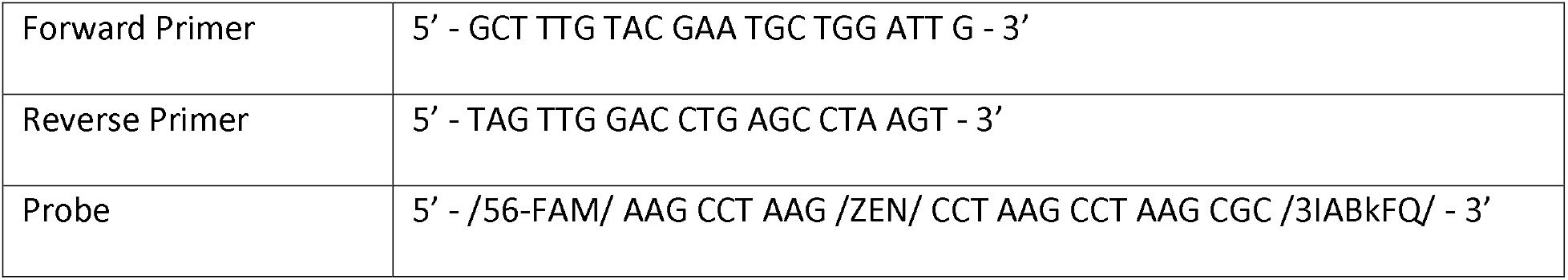
Primer and probe sequences for the *Mp*R1 repeat-targeting index assay.

### Assay Optimization and Validation

#### Primer titrations

Employing titrated primer stocks in a 3 × 3 matrix for testing of gDNA template, optimal concentrations were determined to be 500 pmol/µL for the forward primer and 300 pmol/µL for the reverse primer (S1 Table).

#### Determination of Optimal Reaction Temperatures

By testing optimal primer concentrations (described above) across a gradient of annealing/extension temperatures ranging from 55 °C to 60 °C, advantageous conditions for amplification were determined. As all temperatures resulted in nearly identical Cq values (S2 Table), an annealing/extension temperature of 60 °C was selected in order to minimize risk of non-specific amplification.

#### Examination of Analytical Specificity

Utilizing optimized reaction conditions and cycling protocols, the analytical specificity of our assay was evaluated by testing against human gDNA template, as well as gDNA isolated from the related filarial parasites *B. malayi, B. pahangi, L. loa, A. viteae, O. volvulus, W. bancrofti, D. immitis*, and *M. ozzardi*. No “off target” amplification was observed.

#### Examination of Analytical Sensitivity

Utilizing optimal reaction conditions, all tested samples containing 10 fg or more of DNA template were detected in all replicates.

#### Determination of reaction efficiency

The reaction efficiency was calculated using a titration of our positive control plasmid. Utilizing plasmid size and template mass to estimate target copy number, results were plotted as the log transformed copy number vs. mean Cq value. From the slope of the resulting curve, a reaction efficiency of 97.16% was calculated, along with an amplification factor of 1.97 (Fig 1).

**Figure 1.**
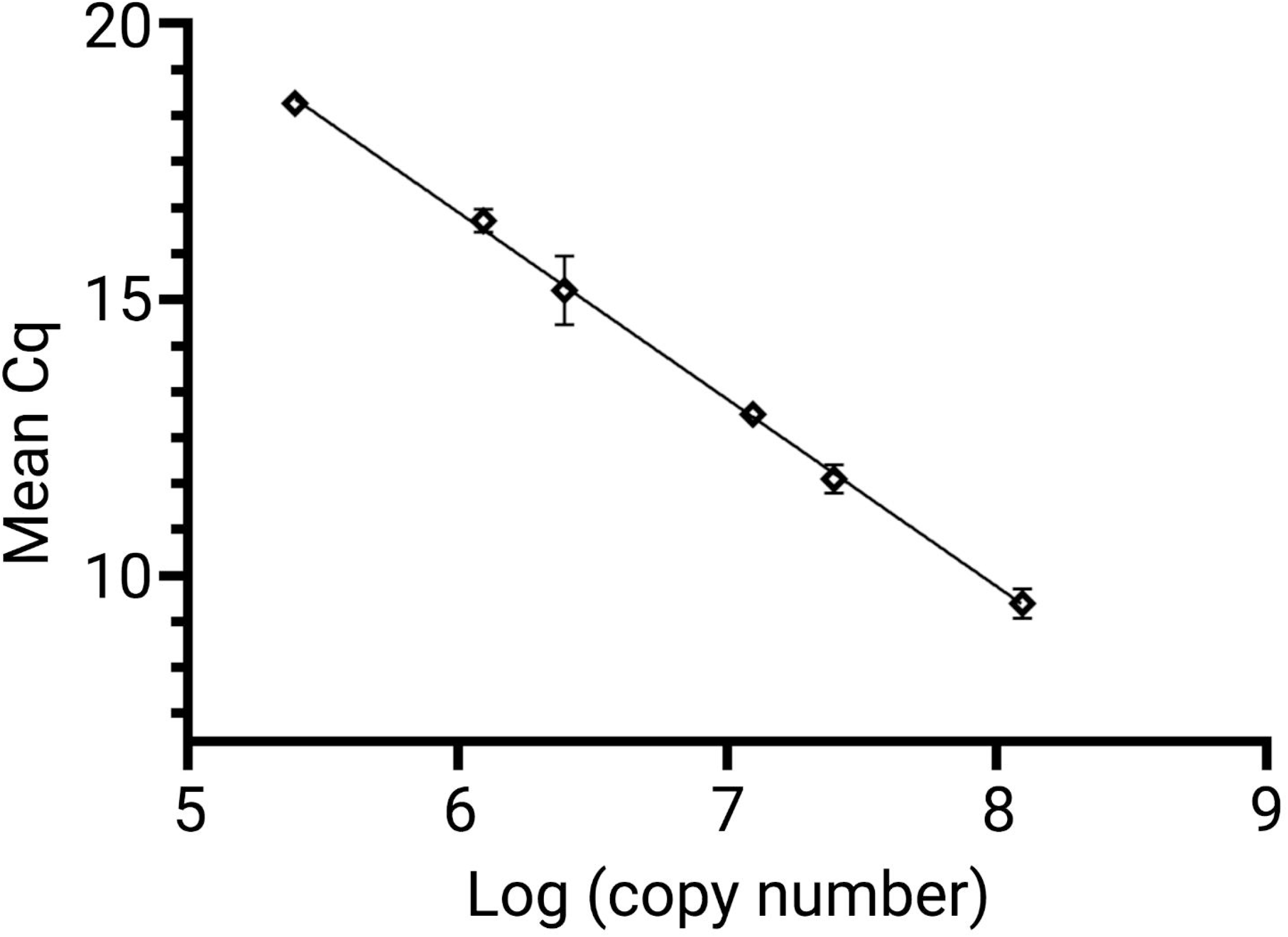
Determination of assay efficiency and amplification factor. To determine assay efficiency and amplification factor, a dilution series was prepared using control plasmid. All dilutions, containing plasmid concentrations ranging from 2.5 × 10^5^ target copies/reaction to 1.25 × 10^8^ target copies/reaction were run in nine to 11 replicate reactions. Mean Cq values and standard deviations were then calculated for all replicates containing each number of template copies, a slope was plotted, and both the reaction efficiency and amplification factor were determined.

### Assessment of Clinical Sensitivity

#### Comparative analysis of field-collected blood samples

Testing of 158 human-blood-derived DNA extracts resulted in the identification of 58 samples (36.7%) which produced a positive qPCR result using both the *Mp*R1 index assay and the previously described ribosomal sequence-targeting reference assay. An additional 95 samples (60.1%) produced a negative result when tested using both assays. Five samples (3.2%) produced a positive result when tested using the *Mp*R1 index assay, but a negative result when tested using the reference assay. No samples were identified as positive using the reference assay, but negative when tested using the *Mp*R1 assay (Table 2, S3 Table). A comparison of Cq values among all co-positive samples resulted in a mean difference of 11.16 cycles, with the *Mp*R1 assay producing a lower mean Cq value for all samples (Fig 2A). A distribution of the by-sample mean Cq value difference across assays is shown in Figure 2B.

**Table 2.**
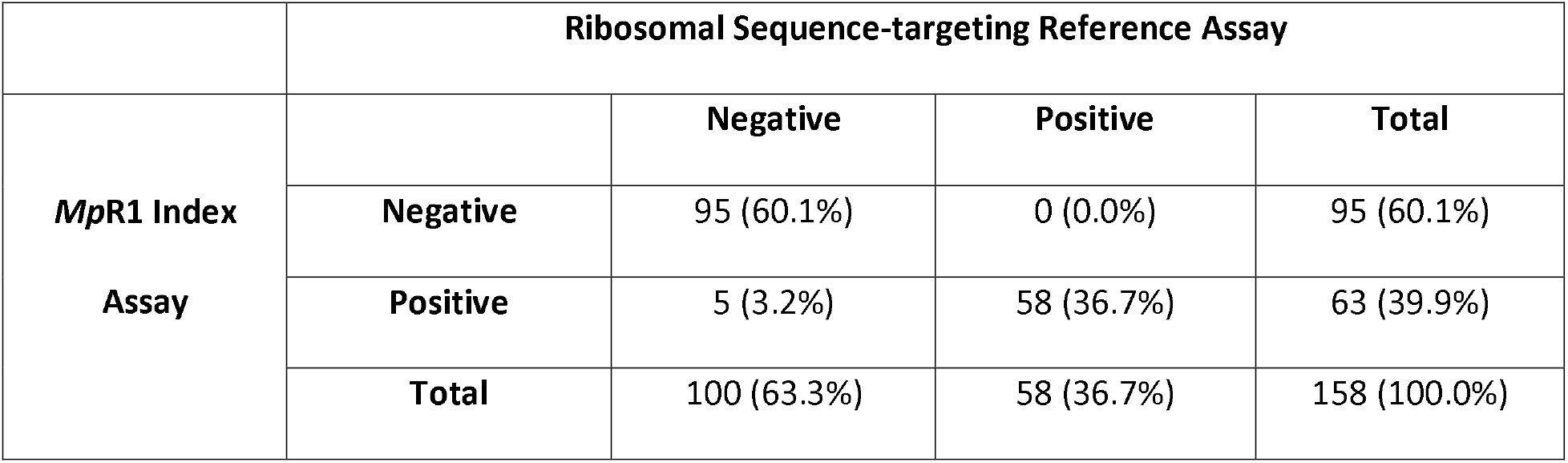
Agreement of assay results upon comparative testing of human bloodspot samples.

**Figure 2.**
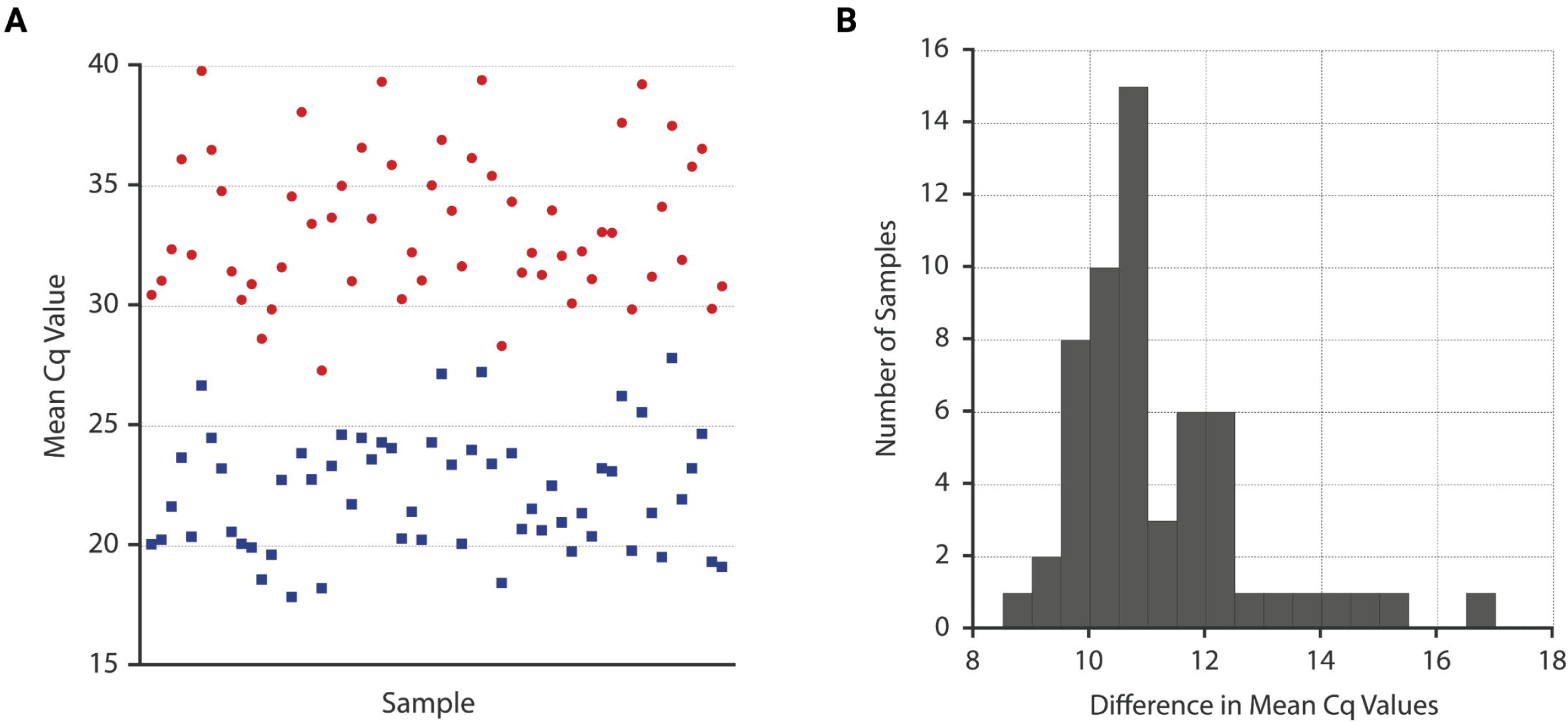
Differences between mean Cq values for samples producing positive results for both reference and index assays. **(A)** For each co-positive sample, mean Cq values were plotted for both the ribosomal ITS-targeting reference assay (red circles) and the repetitive sequence-targeting (*Mp*R1) index assay (blue squares). **(B)** For all co-positive samples, a difference was calculated by subtracting the mean Cq value for the results of index assay testing from the mean Cq value resulting from reference assay testing. Differences were then binned and plotted. The average difference resulting from all plotted (co-positive) samples was 11.16 cycles.

#### Comparative analysis of field-collected mosquito samples

A total of 316 DNA samples, isolated from individual mosquito carcasses, were tested for the presence of *M. perstans* using both index and reference assays. Following initial testing, 25 samples were eliminated from the data set due to an insufficient sample volume, preventing the retesting of inconsistently positive results from one, or both assays. Of the remaining 291 samples, 262 extracts (90.0%) were determined to be negative using both assays, with 16 samples (5.5%) testing positive for *M. perstans* when targeting both index and reference targets. An additional 13 samples (4.5%) produced positive results when testing for the *Mp*R1 assay’s repetitive target, but negative results when assaying for the ribosomal repeat. Not a single sample produced a reference assay positive result coupled with an index assay negative result (Table 3, S4 Table).

**Table 3.** Agreement of assay results upon comparative testing of field-collected mosquito samples.

|  | Ribosomal Sequence-targeting Reference Assay |  |  |  |
| --- | --- | --- | --- | --- |
| *MpR1* Index<br>Assay |  | Negative | Positive | Total |
|  | Negative | 262 (90.0%) | 0 (0.0%) | 262 (90.0%) |
|  | Positive | 13 (4.5%) | 16 (5.5%) | 29 (10.0%) |
|  | Total | 275 (94.5%) | 16 (5.5%) | 291 (100%) |

## Discussion

Spanning multiple continents and vast geographic regions, *M. perstans* is an understudied pathogen whose importance for human health remains poorly understood. Yet despite a historical lack of research interest, a trend towards further exploration of the health impacts of mansonellosis has recently emerged, with growing evidence suggesting that infection outcomes may result in greater morbidity than previously believed [4]. To properly evaluate the impact and potential of such concerns, additional diagnostic research tools, capable of sensitively and specifically identifying positive samples will be of critical importance.

Attempting to meet this need, we have identified a high copy-number repetitive sequence which we have exploited as a qPCR assay target. Testing this *Mp*R1 assay against a ribosomal sequence-targeting reference assay [5,26], our index assay demonstrated improved analytical sensitivity, facilitating consistent detection of as little as 10 fg of pathogen-derived gDNA. Moreover, clinical sensitivity was also improved, with additional positive samples identified when testing human blood-derived DNA extracts (Table 2). Of note, while a poor measure of sensitivity, a comparison of Cq values can provide insight into the relative prevalence of target sequences between assays with similar efficiencies. As such, an examination of comparative Cq values suggested a higher prevalence of the index assay’s repetitive target than the employed reference assay’s ribosomal sequence target, with a mean difference in Cq values of 11.16 cycles. Such improvements to both analytical and clinical sensitivity will provide programmatic decision makers with a tool capable of producing an accurate picture of infection status, should the growing momentum for the expanded study of *M. perstans* result in tangible operational research efforts.

While the testing of human blood samples remains the most obvious application of our novel assay, a demonstrated ability to test mosquito samples for the presence of *M. perstans* is also noteworthy. This capability not only has the potential to facilitate mapping and surveillance efforts by serving as a non-invasive proxy for human infection, but it also has the capacity to facilitate studies of vector incrimination. As previous studies have hypothesized the potential existence of a *M. perstans* mosquito vector in select geographic settings [2], the testing of mosquitoes will allow for expanded insight into such a possibility. Penetration of the mosquito midgut and partial development of *Mansonella ozzardi* has already been documented in *Aedes aegypti* [32], warranting the assessment of any similar abilities in possible *M. perstans* vectors. Accordingly, insights into partial, or complete pathogen development could be garnered through the independent testing of mosquito carcass sections (abdomen, head, and thorax) to examine worm migration patterns within the mosquito host. However, even if mosquito vectoring capacity is limited or nonexistent, an improved understanding of the behavior and/or partial development of *M. perstans* within the mosquito host would be valuable, as midgut penetration by filarial parasites has been shown to aid transmission of viral pathogens, including Dengue and Eastern Equine Encephalitis, under conditions of co-infection [33,34]. As such, an improved understanding of *M. perstans* development within the mosquito could provide valuable information for modeling efforts of viral transmission in co-endemic settings.

Given the significant number of open research questions that pertain to the *M. perstans-* mosquito relationship, an ability to analyze mosquitoes for the presence of this pathogen provides excellent opportunities for the integration of surveillance, allowing mosquitoes collected for other purposes to be screened for the presence of this “non-vectored” pathogen. Through the creative and expanded use of such captured mosquito populations, these efforts should allow study planners to develop useful collaborative partnerships. Such partnerships would improve the overall return on research investments and maximizing the volume of data collected from each effort.

One noted shortcoming of this study has been an inability to source, and therefore validate our assay against DNA isolated from *Mansonella streptocerca*. As *M. streptocerca* and *M. perstans* overlap geographically [3,35], and ribosomal sequence analysis has suggested a closer phylogenetic relationship between *M. perstans* and *M. streptocerca* than between *M. perstans* and *M. ozzardi* [3], testing against this closely related parasite will be important to fully validate our assay’s capacity to discriminate human-infecting pathogens of the *Manonella* genus to the species level. Efforts to identify a source for *M. streptocerca* material, and other *Mansonella* spp. such as *Mansonella rodhaini* are ongoing, as are plans for additional validation efforts.

Through the systematic identification of an optimal DNA-based diagnostic target, we have developed a qPCR assay with the capacity to improve detection of *M. perstans* in both human and mosquito samples. This assay provides a useful tool for the potential expansion of *M. perstans* monitoring efforts and also serves to facilitate integrated xenosurveillance efforts. Given the lack of knowledge surrounding the distribution, parasite-vector relationships, and clinical impact of this parasite, tools capable of providing improved insight into its prevalence should facilitate expanded study of this parasite within the research community.

## Supporting information

S1 Checklist

## Data Availability

All data produced in the present study are available upon reasonable request to the authors.

## Acknowledgements

The authors are indebted to Edward J. Tettevi, Francis B. D. Veriegh, and Mike Yaw Osei-Atweneboana for their involvement in the primary study that provided the research resources for the validation of this assay. Computational resources were provided by the ELIXIR-CZ project (LM2015047), part of the international ELIXIR infrastructure.

## Supporting Information Legends

**S1 Checklist. STARD checklist.** Locations within the manuscript at which each checklist item is addressed are identified. This checklist is included to facilitate the reader’s ability to assess potential study biases and to facilitate consideration of the generalizability of reported results.

**S1 Table. Results of primer titration experiments.** Cq values resulting from reactions performed as part of primer titration experiments are provided.

**S2 Table. Results of temperature optimization experiments.** Cq values resulting from reactions performed as part of temperature optimization experiments are provided.

**S3 Table. Individual reaction results of field-collected blood sample testing.** Cq values resulting from the testing of all blood samples are provided for both reference and index assays.

**S4 Table. Individual reaction results of field-collected mosquito sample testing.** Cq values resulting from the testing of all mosquito samples are provided for both reference and index assays.

